# National- and state-level SARS-CoV-2 immunity trends from January 2020 to December 2023: a mathematical modeling analysis

**DOI:** 10.1101/2024.10.22.24315935

**Authors:** Fayette Klaassen, Nicole A Swartwood, Melanie H Chitwood, Rafael Lopes, Masahiko Haraguchi, Joshua A Salomon, Ted Cohen, Nicolas A Menzies

## Abstract

**Introduction:** Effective immune protection against SARS-CoV-2 infection and severe COVID-19 disease continues to change due to viral evolution and waning immunity. We estimated population-level immunity to SARS-CoV-2 for each of the fifty United States (U.S.) and the District of Columbia from January 2020 through December 2023.

**Methods:** We updated a model of SARS-CoV-2 infections to align with the latest evidence on SARS-CoV-2 natural history and waning of immunity, and to integrate various data sources available throughout the pandemic. We used this model to produce population estimates of effective protection against SARS-CoV-2 infection and severe COVID-19 disease.

**Results:** On December 30, 2023, 99.9% of the U.S. population had experienced immunological exposure to SARS-CoV-2 through infection and/or vaccination, with 99.4% (95% credible interval (CrI): 92.4-100%) having had at least one SARS-CoV-2 infection. Despite this high exposure, the average population-level protection against infection was 53.6% (95% CrI: 38.7-71.5%). Population-level protection against severe disease was 82.6% (95% CrI: 71.5-91.7%).

**Discussion:** A new wave of SARS-CoV-2 infections and COVID-19-associated hospitalizations began near the end of 2023, with the introduction of the JN.1 variant. This upturn suggests that the U.S. population remains at risk of SARS-CoV-2 infection and severe COVID-19 disease despite the high level of cumulative exposure in the United States. This decline in effective protection is likely due to both waning and continued viral evolution.

## Introduction

On May 11, 2023, the United States (U.S.) ended the COVID-19 U.S. Public Health Emergency [1], resulting in the dissolution of many surveillance systems developed to track severe acute respiratory syndrome coronavirus 2 (SARS-CoV-2)[2], the removal of remaining non-pharmaceutical protections against SARS-CoV-2 transmission, and the cessation of free testing, vaccination, and treatment services [3]. However, SARS-CoV-2 continues to evolve. New variants—including JN.1, which emerged in late 2023—have increased COVID-19 cases and associated hospitalizations [4, 5].

At the population level, effective immunological protection has varied over the course of the pandemic, with each successive SARS-CoV-2 variant bringing risks of immune escape, most notably following the introduction of the BA.1 (Omicron) variant in late 2021 [6]. Additionally, previously acquired immunological protection via vaccination or SARS-CoV-2 infection wanes over time [7–9]. In response to the emergence of new variants and the waning of immunity, updated vaccines designed to match circulating variants have been introduced to attempt to maintain high levels of population protection [10].

The dynamics of population-level immunity, affected by both the emergence of variants and waning immunity after vaccination or natural infections, greatly affect the risk of viral emergence and spread. Immunity conferred by COVID-19 infection and vaccination also provides protection against the development of severe COVID-19 disease [11, 12] and persistent, chronic symptoms after resolution of acute SARS-CoV-2 infection (long COVID) [13]. Despite the continued importance of population-level immunity, the most recent time series of immunity estimates ended in November 2022 [14]. Furthermore, the previously available time series separated immunity estimates into pre-Omicron and Omicron-specific trends [14, 15]. In this analysis, we estimated population-level immunity to SARS-CoV-2 for each of the fifty U.S. states and the District of Columbia from January 2020 through December 2023. To do so, we updated a model of SARS-CoV-2 infections to incorporate the latest understanding of the virus’s natural history and waning immunity and to integrate various data sources available throughout the pandemic. We used this model to produce estimates of population effective protection from SARS-CoV-2 infection and severe COVID-19 disease.

## Methods

### Data

We used daily reported COVID-19 case and COVID-19 death counts compiled by the Johns Hopkins Center for Systems Science and Engineering [16], available from the first reported case [range: January 13, 2020 (Washington) – March 12, 2020 (Colorado)] through March 9, 2023. We extended these case data with case reports from the CDC [17], available through May 11, 2023 (March 30, 2023, for Iowa). We extracted hospitalization records from the U.S. Health and Human Services [18], which were available from the first hospitalization report [range: July 25, 2020 (Connecticut and Nevada) – October 17, 2020 (Texas and North Dakota)] through April 21, 2024. We extracted vaccination records reflecting the first vaccination series (“received first vaccination”) and boosters (“received booster dose”) from the CDC [19]. These data are available from February 13, 2021, through May 10, 2023. The date of first reported vaccination series and booster dose is lagged compared to the start of the vaccination roll-out. Therefore, we distributed the first vaccination series/booster entry linearly over the six preceding weeks. We extended the booster data with monthly coverage estimates of the updated 2023-2024 COVID-19 vaccine from the National Center for Immunization and Respiratory Diseases (NCIRD) [20], available from September 2023 through August 2024. For seven states (Kansas, Massachusetts, Nebraska, New Hampshire, Rhode Island, South Carolina, and Virginia), monthly NCIRD reports were unavailable, and weekly estimates from the CDC’s National Immunization Survey were used instead [21] (Supplementary Methods). All data were aggregated into weekly counts for each of the fifty U.S. states and the District of Columbia.

### Immunological exposure states

We calculated the percentage of the population who were immunologically exposed (through either infection or vaccination) by week and U.S. state. Immunological exposure was categorized as infected (never vaccinated), vaccinated (never infected), and hybrid (at least one infection and at least one vaccination series/booster) (Figure 1A). Exposure states were calculated at each time point using the vaccination and booster data, as well as infection estimates. New infections were distributed over exposure states proportional to the effective protection for each state. Following Klaassen et al. (2022) [15], we estimated hybrid immunity (i.e., immunity achieved through both infection and vaccination) using an odds ratio for vaccine receipt among individuals with versus without prior infection, based on data collected in the U.S. Census Bureau Household Pulse Survey [22]. Using this odds ratio, initial vaccinations and subsequent boosters were assigned to the applicable exposure states. Initial vaccination series could only be administered in the naïve and infected exposure states; boosters could only be administered in the vaccinated and hybrid exposure states (Supplementary Methods).

**Figure 1:**
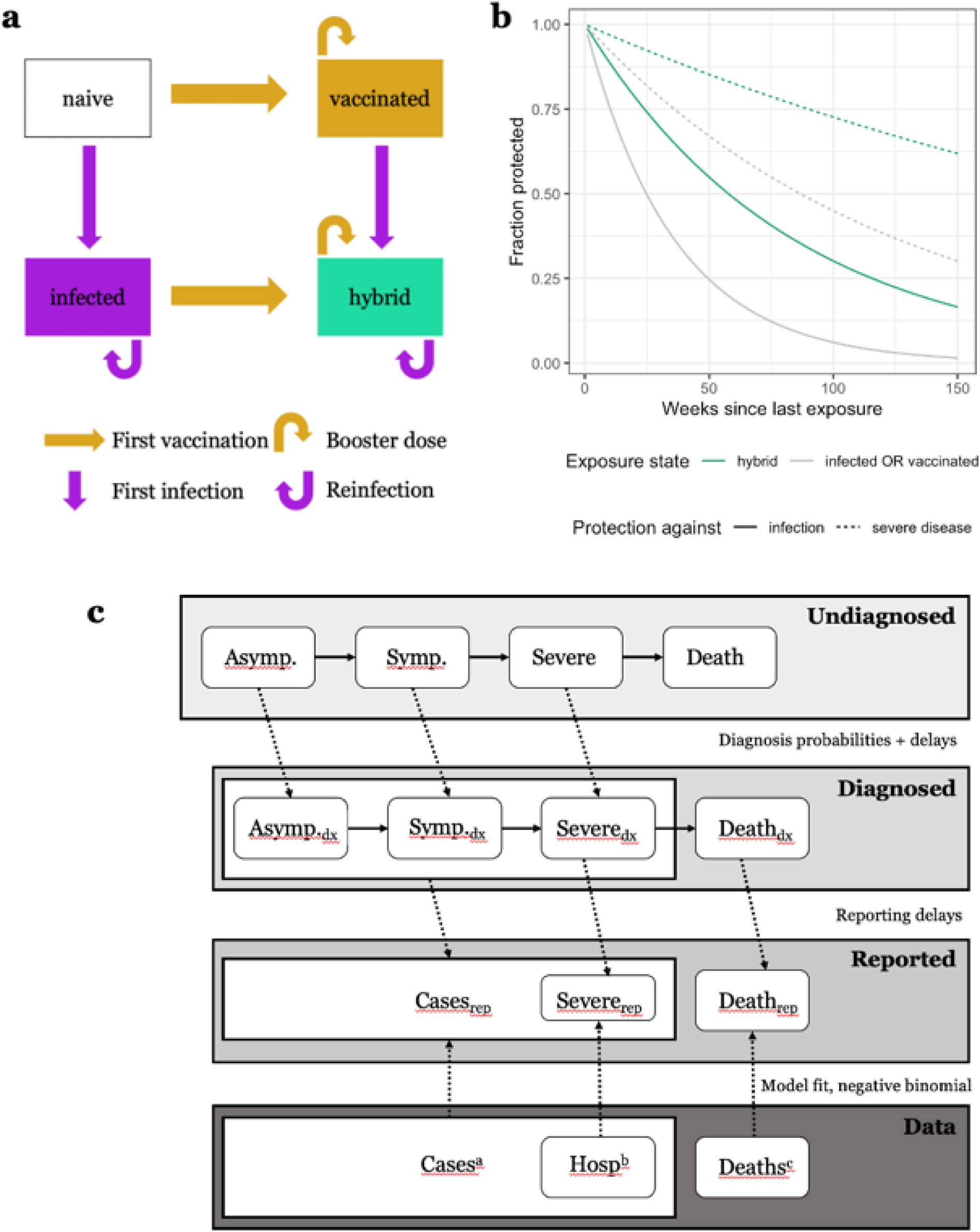
Key model assumptions and states. Panel A: Immune exposure state diagram. Panel B: Immunity waning assumptions. Panel C: Model state diagram. Horizontal arrows indicate progression probabilities and delay. Vertical arrows indicate data fit to model. Diagonal arrows indicate delays and probabilities from undiagnosed disease state to diagnosed and reported states. Not pictured: at any disease state (other than death), there is a probability of recovery. Definition of abbreviations in cells: *Asymp*. = asymptomatic disease, *Symp*. = symptomatic disease, *Severe* = severe disease, *Hosp* = hospitalized. Definition of subscripts: *dx* is diagnosed, *rep* is reported. Notes: *a*. Case data are available from start date through May 11, 2023; *b*. hospitalization data are available from August 2020 through the end of the time series; *c*. death reports are available from the start date through March 2023.

### Waning of protection

Each exposure state was assumed to confer protection against both infection and severe disease, with the level of protection differing between these outcomes and declining over time. The *naïve* population was assumed to be fully susceptible to infection and subsequent severe disease. Informed by clinical trials, antibody titers, and vaccination effectiveness studies [23–27], we formulated assumptions for the waning of protection against infection and severe disease, stratified by exposure state (Figure 1B). Antibody titers are believed to wane at similar rates among infected and vaccinated persons [27], and we applied the same waning assumptions to the *infected* and *vaccinated* populations, consistent with the approach in Klaassen et al. 2022 [15]. We assumed protection against infection is 80% immediately following first exposure and wanes exponentially, with a weekly rate of 2.8% (50% of protection lost by 25 weeks). For these individuals, we assumed that protection against developing severe disease after a subsequent infection starts at 95% and wanes exponentially at a rate of 0.8% (50% of protection lost by 86 weeks). Immunity through infection and vaccination (*hybrid)* immunity is more durable than immunity through infection or vaccination alone [26]. Therefore, for the *hybrid* exposure state, we assumed a slower waning of immunological protection, consistent with Klaassen et al. 2023 [14]. We assumed protection against infection is 80% following new exposure and wanes exponentially at a rate of 1.2% (50% of protection lost by 58 weeks), and that protection against developing severe disease after a subsequent infection starts at 95% and wanes exponentially with a rate of 0.3% (50% of protection lost by 217 weeks). Finally, we assumed that when the Omicron variant emerged (December 1, 2021), both the previously acquired protection against infection and the protection against severe disease were reduced by 70%, consistent with previous model assumptions to account for immune escape [15].

### Mathematical model

We updated *covidestim,* a previously published mathematical model of SARS-CoV-2 infections in the United States [14, 15, 28]. The model uses a mechanistic back-calculation approach to estimate infections and subsequent health states based on observed COVID-19 cases, COVID-19-associated hospitalizations, COVID-19-associated deaths, and COVID-19 vaccination data. In this model, probabilities of and delays in progression between disease states were informed by published estimates [29–37], and partial protection conferred by infection and/or vaccination was explicitly encoded (Figure 1C). The model used a weekly timestep to match the reporting interval of hospitalization data. Further details on the model can be found in the Supplementary Methods and Supplementary Table 1.

#### Disease progression

We adjusted progression probabilities to account for protection against severe disease among persons who had a prior SARS-CoV-2 exposure. Immunological protection was assumed to decrease the probabilities of progressing from infected to symptomatic states and from symptomatic to severe states. (Supplementary Table 2 and Supplementary Methods).

#### Transition to Omicron dominance

We assumed that the transition to Omicron-variant dominance occurred during December 2021. We implemented a linear transition of variant-specific parameters between December 1, 2021, and January 1, 2022. We also assumed a 70% reduction in immunity for individuals immunologically exposed to pre-Omicron variants. Supplementary Table 2 lists these variant-specific parameters and their priors.

## Statistical analysis

We estimated the model using a Bayesian statistical approach (a Hamiltonian Monte Carlo sampling algorithm, as implemented by the Stan software library [38]). We ran the model for each U.S. state using 3 chains, 1000 iterations each, with 2/3 of these iterations devoted to warmup, rendering a total of 1000 sampled parameter sets. For state-level estimates, we report uncertainty as equal-tailed 95% credible intervals. For the U.S. Census region and U.S. national estimates, we calculated the point estimate as the sum of the state-level estimates and intervals as the sum of the lower and upper bounds of state-level intervals. Vermont was excluded from the state-level and aggregated (region and national) results due to model non-convergence. All analyses were run in R (version 4.2.1) using the *rstan* and *covidestim* packages [39–41]. Model output is available on Harvard Dataverse by state and as summary files [42].

## Results

### Model performance

Figure 2 shows the model fit to COVID-19 cases, COVID-19–associated deaths, and COVID-19–associated hospitalizations, compared with available data in California, North Dakota, Georgia, and Massachusetts (one state from each of the four U.S. Census regions) during 2020– 2023 (Supplementary Figure 1 shows model performance for each of the fifty states and District of Columbia over 2020–2023). The model fitted better to the hospitalization data than to the COVID-19 death data. Figure 2 also shows the model-estimated SARS-CoV-2 infections for each of the four states.

**Figure 2:**
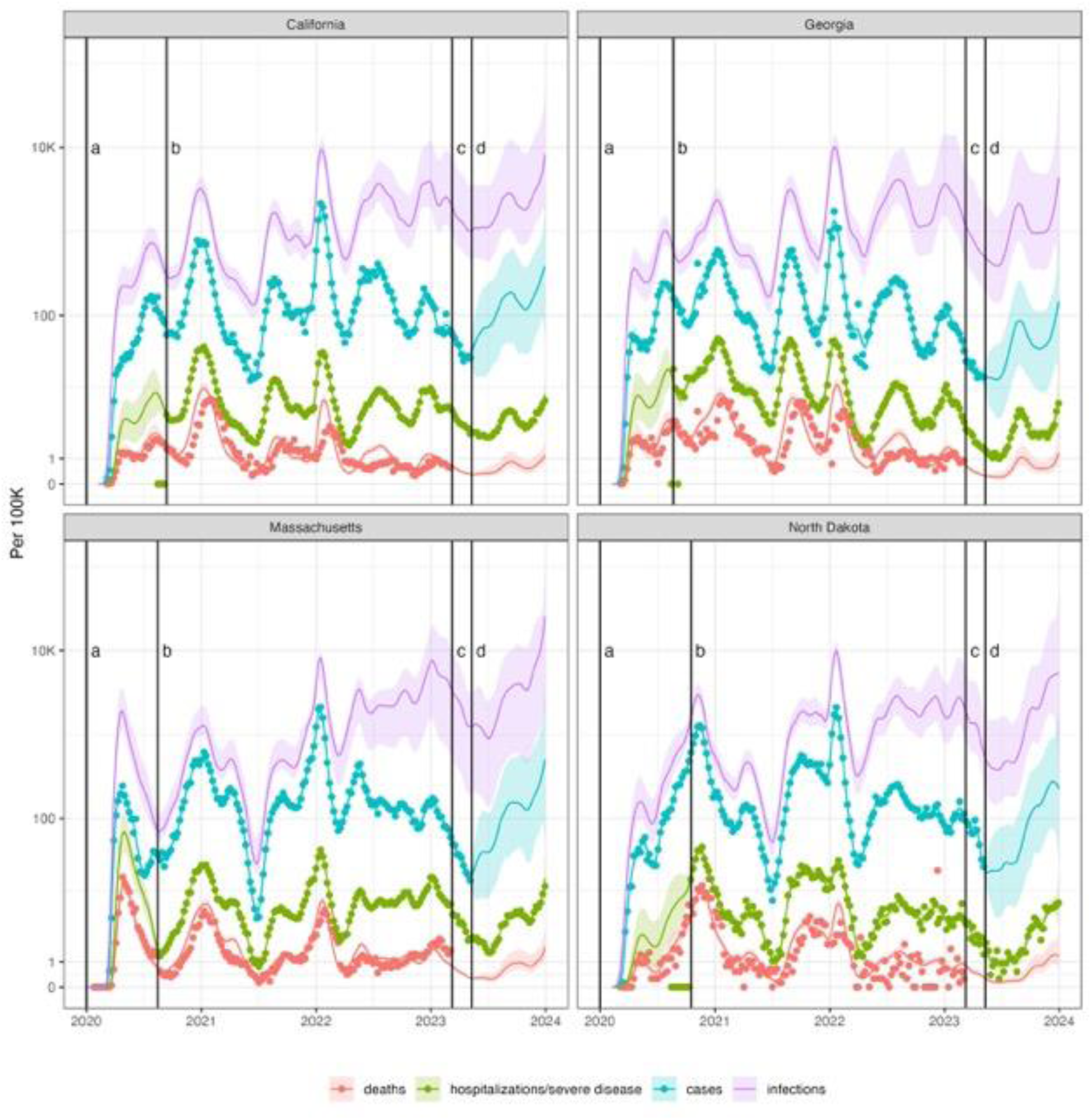
Model estimated trend in SARS-CoV-2 infections, total COVID-19 cases, severe COVID-19 cases, and deaths with COVID-19 compared with available reported data for the United States and four selected states for 2020-2023. Note: Time range ‘a’ represents model fits using only COVID-19 cases and deaths data. Time range ‘b’ represents model fits using COVID-19 cases, deaths, and hospitalizations data. Time range ‘c’ represents fits using only COVID-19 cases and hospitalizations data. Time range ‘d’ represents model fit using only COVID-19 hospitalizations data.

### Immunity estimates

All Census regions exceeded 95% of the population immunologically exposed over the course of 2022 (dates of reaching these thresholds by region: Northeast: January 8, 2022; West: January 29, 2022; South: February 5, 2022; Midwest: April 30, 2022). Figure 3 depicts cumulative immunity exposure (infected, vaccinated, and hybrid) for the United States and four Census regions from the beginning of 2020 to the end of 2023. On December 30, 2023, 99.4% [92.4%, 100%] of persons in the United States were estimated to have had at least one SARS-CoV-2 infection. The average protection in the United States against severe disease was 82.6% [95% credible interval (CrI) 71.5%, 91.7%] and against infection 53.7% [95% CrI 38.6%, 71.5%]. On this date, population-level estimates of protection against severe disease across states ranged from 71.1% [59.3%, 87.0%] in Mississippi to 90.5% [80.8%, 93.2%] in Maine. Estimates of protection against infection ranged from 36.1% [24.8%, 60.7%] in Mississippi to 67.7% [49.5%, 75.0%] in Maine. Supplementary Figure 2 shows exposure states and protection over time for each of the fifty U.S. states and the District of Columbia.

**Figure 3:**
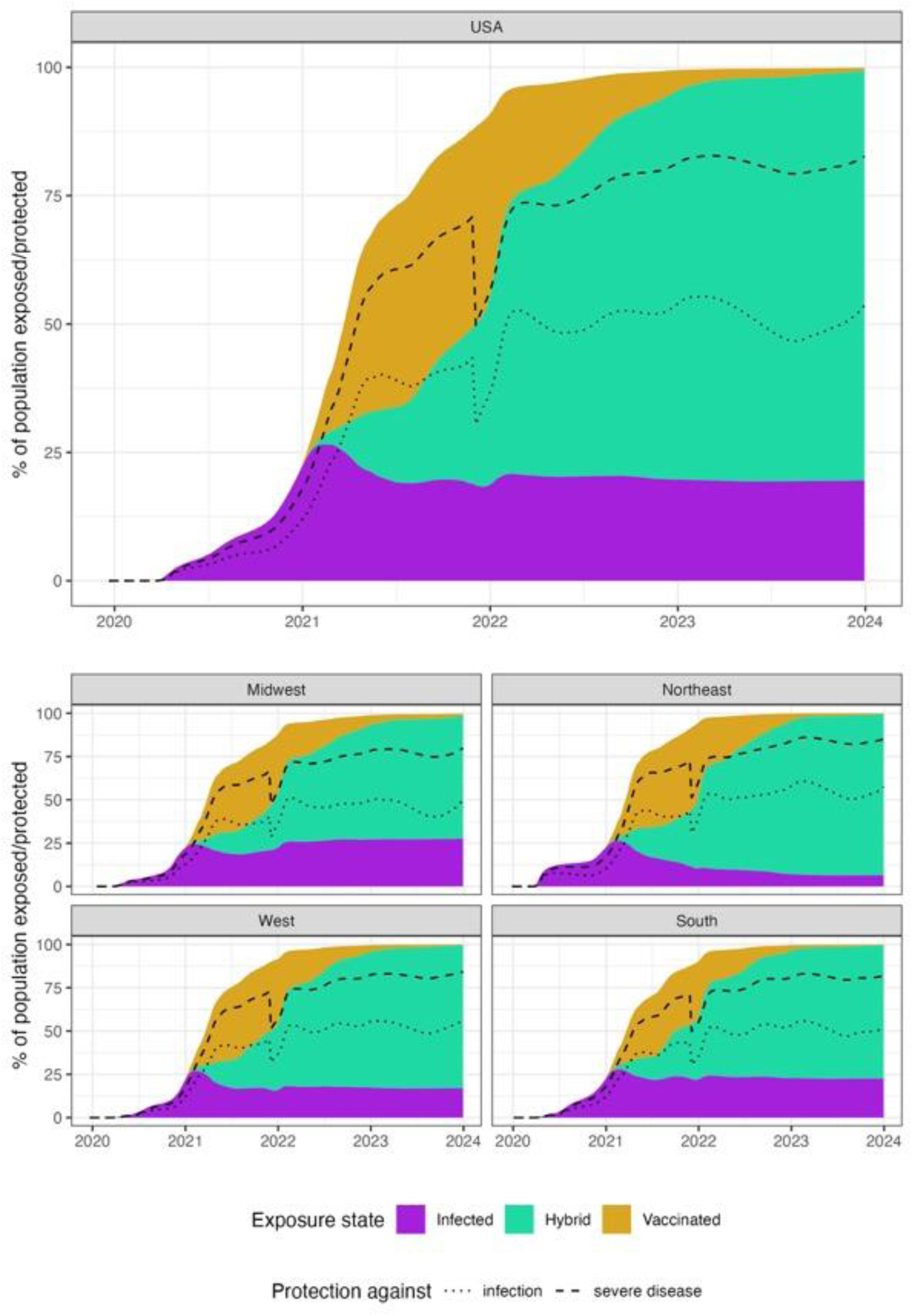
Percentage of population immunological exposed and protected against SARS-CoV-2 infection and severe COVID-19 disease stratified exposure route and U.S. Census Region.

The largest loss of immunological protection occurred in December 2021, largely due to the immune escape associated with the Omicron variant (Northeast: 18.4% [17.2%, 19.9%]; West: 17.2% [16.0%, 18.7%]; South: 17.2% [15.7, 19.0%]; Midwest: 16.0% [14.6%, 17.9%]). Figure 4 shows the 2020–2023 time-trends in the population percentage in immunity against SARS-CoV-2 infection for the United States and the four U.S. Census regions. Supplementary Figure 3 shows the trends in effective protection for each of the fifty U.S. states and the District of Columbia.

**Figure 4:**
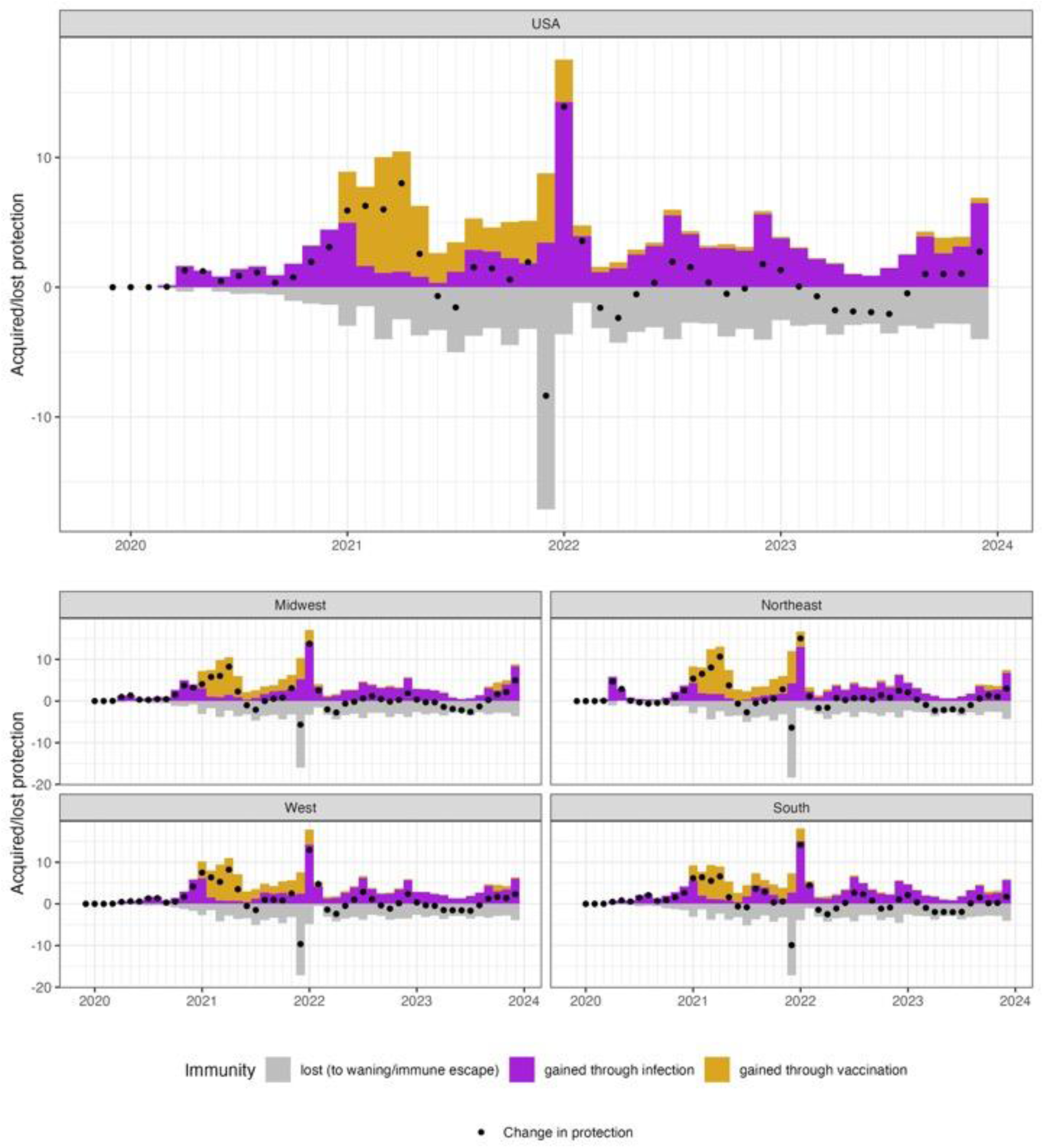
Monthly change in acquired immunity stratified by exposure route and U.S. Census Region, 2020–2023

Figure 5 summarizes the relationship between the total protection gained in the population from SARS-CoV-2 infection and COVID-19 vaccination for each of the fifty U.S. states and the District of Columbia. This outcome only reflects the increase in effective protection and is lower than total immunological exposures. This difference is because a repeat exposure only increased effective protection by the amount corresponding to the difference between full protection and current protection retained from a previous exposure. While the total protection gained over the course of the pandemic was similar across states (range: 164% [120%, 233%] for the District of Columbia; to 213% [176%, 268%] for New Mexico), there was a wide range in the fraction of total gained protection from infection, ranging from 0.59 [0.43, 0.63] in District of Columbia (78.9% [69.0%, 85.5%] from vaccination versus 85.2% [51.4%, 148%] from infection) to 0.77 [0.70, 0.82] in Alabama (45.7% [39.7%, 51.2%] from vaccination versus 153% [90.6%, 237%] from infection).

**Figure 5:**
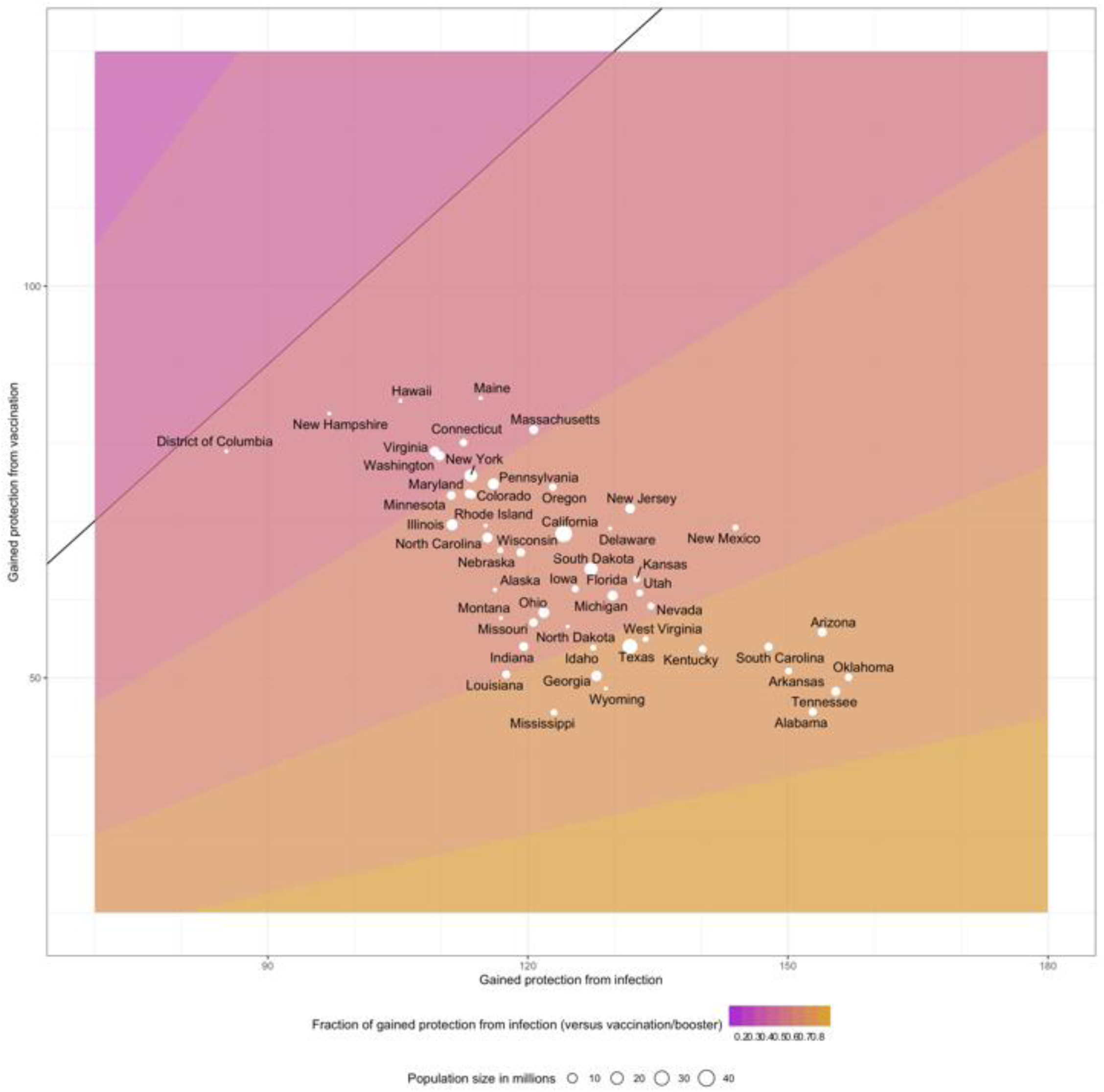
Fraction of total gained effective immunological protection from vaccination and infection by U.S. state, 2020–2023

## Discussion

The combined effect of viral evolution and waning of acquired immunity has resulted in dynamic changes in effective immunological protection against SARS-CoV-2 infection in the United States. In this model-based analysis, we quantified population-level protection against SARS-CoV-2 infection and severe COVID-19 disease for each U.S. state and the District of Columbia from January 2020–December 2023. We estimated that 99.9% [99.2%, 100%] of the U.S. population had been exposed to SARS-CoV-2, either through infection or vaccination, by December 2023. Despite near-universal exposure, only 53.6% [38.7%, 71.5%] of persons retained effective protection against SARS-CoV-2 infection at that time, a level only marginally higher than the lowest levels observed in the post-Omicron period (46.7% [34.9%, 59.7%], on August 19, 2023).

Immunological exposure and effective protection varied across the United States. Despite reaching similar levels of total protection against infection as other regions, the Northeast had lower rates of infection-only exposure and subsequent protection during 2021–2023, reflecting consistently high vaccination rates [21]. In the Northeast, only 3.2% [1.6%, 4.9%] had protection against infection because of infection-only immunological-exposure; the only infected exposure protection was considerably larger in the West (8.3% [4.7%, 12.5%]), the South (10.0% [5.5%, 15.8%]) and the Midwest (11.9% [6.4%, 20.0%]). The trends in these immunity estimates reflect the different drivers of population protection against SARS-CoV-2 infections. In 2020, SARS-CoV-2 was introduced into a naïve population, and immunity was gained only through infections. In early 2021, immunity increased rapidly due to vaccine introduction. These gains were the smallest in the South, which experienced relatively high rates of vaccine hesitancy [43]. Despite these gains in immunity, the emergence of the Omicron variant and its associated immune escape contributed to the most significant decrease in immunity between 2020 and 2023.

Descendants of initial Omicron lineages have also been able to evade immunity to earlier variants. However, this effect has diminished as more individuals have been infected by Omicron sub-lineages. In December 2023, there was a 7% increase in immunity due to infection, making the largest single-month increase in immunity since the Omicron takeover. This increase in infection-acquired immunity is likely due to the introduction of the JN.1 Omicron variant, which quickly comprised up to 29% of SARS-CoV-2 infections in the U.S. by the end of 2023 [4]. In addition to the characteristic Omicron immune escape of its parental lineage BA.2.86.1, the JN.1 variant gained its further advantage through additional mutations, which have likely increased transmissibility [5]. Continued viral evolution and waning immunity suggest that a recurrent vaccination strategy as well as recurrent updating of vaccine formulation may be needed to maintain effective protection against SARS-CoV-2 infection.

We note that the availability of COVID-19 case, death, hospitalization and vaccination data in the U.S. varied over the study period. Here, we aimed to include all available data and bridge any data gaps to provide the model with as much evidence as possible. Accordingly, we have higher confidence in the trends in SARS-CoV-2 infections and immunity than previously presented estimates [14, 15], as the underlying model has been retrospectively fit to all identified data sources (Supplementary Figure S4).

Our study had several limitations. First, the vaccination datasets used in the analysis have associated biases. The initial dataset reported by the CDC is known to be nonrepresentative for some locations [44]. While a potential underreporting of vaccination might result in an underestimation of immunity, this bias is likely small due to the high level of exposure via infection. The additional NCIRD data reporting vaccination coverage for the second half of 2023 may suffer from the same bias as the initial vaccination dataset. NCIRD data was not available for all states. For the seven states that did not have NCIRD data, we used estimates from an interview-based survey. Survey data may carry biases such as recall and selection biases.

Second, we assumed the level of immunity gained through SARS-Cov-2 infection was constant across all pre-Omicron variants and all Omicron sub-lineages, respectively. We also assumed the same immunity for different vaccine products. Furthermore, we assumed an accelerated waning of immunity for a single exposure (i.e. infection or vaccination) compared with hybrid exposure. This differential waning assumption results in higher immunity than if hybrid waning was equal to that of single exposure (Supplementary Figure S5).

By the end of 2023, nearly the entire U.S. population had been exposed to SARS-CoV-2 through either infection or vaccination. However, due to the continued evolution of the virus and waning immune protection, only 56% of persons retained effective protection against infection, and 81% retained protection against severe COVID-19 disease. Due to this declining protection, the beginnings of a new wave of SARS-CoV-2 infections and COVID-19-associated hospitalizations emerged at the end of 2023, with the introduction of the JN.1 variant. This upturn suggests that the U.S. population remains at risk of SARS-CoV-2 infection and severe COVID-19 disease despite the high level of cumulative exposure in the United States.

## Data Availability

All code and data used in this analysis are available on GitHub (https://github.com/covidestim). The produced datasets are available on the Harvard Dataverse (https://doi.org/10.7910/DVN/G2ZXJG).

## Acknowledgements

We thank Stephanie Su for assistance in study coordination.

## Author Contributions

F.K. and N.A.S. conceived the project. T.C., J.A.S. and N.A.M. acquired funding. F.K. wrote the model code, curated the data and executed the analysis, and visualized the results. N.A.S. and F.K. drafted the initial manuscript. All authors contributed to the development of the methodology, reviewed, and edited the original manuscript.

## Disclaimer

The findings, conclusions, and views expressed are those of the authors and do not necessarily represent the official position of the Centers for Disease Control and Prevention (CDC), Council of State and Territorial Epidemiologists, or the National Institutes of Health. The funders had no role in study design, data collection and analysis, decision to publish, or preparation of the manuscript.

## Data sharing statement

All code and data used in this analysis are available on GitHub (https://github.com/covidestim). The produced datasets are available on the Harvard Dataverse (https://doi.org/0.7910/DVN/G2ZXJG).

