## Supplementary for "National- and state-level SARS-CoV-2 immunity trends from January 2020 to December 2023: a mathematical modeling analysis"

**Contents**

**Supplementary methods**

I. Modeled disease states
II. Progression probabilities and adjustment
III. Calculation of exposure state transitions and protection
IV. Vaccination data smoothing
V. Table S1: *covidestim* model parameters.
VI. Table S2: Additional *covidestim* model parameters and prior distributions.

**Supplementary results**

I. Table S3: Summary of immunological protection for the United States, U.S. Census Regions, U.S. States and the District of Columbia, 2020-2023.
II. Figure S1: Model estimated SARS-CoV-2 infections and COVID-19 disease outcomes compared with reported data, for each U.S. state and District of Columbia, January 2020–December 2023.
III. Figure S2: Percentage of population immunological exposed and protected against SARS-CoV-2 infection and severe COVID-19 disease stratified exposure route and U.S. state, January 2020 – December 2023.
IV. Figure S3: Monthly change in acquired immunity stratified by exposure route and U.S. state, 2020–2023.
V. Figure S4: Comparison of SARS-CoV-2 infection estimates updated *covidestim* model versus earlier divided pre- and post-Omicron model estimates.
VI. Figure S5: Comparison of effective protection against SARS-CoV-2 infection and severe COVID-19 disease with and without hybrid immunity specific waning assumptions, for each U.S. state and the District of Columbia, January 2020–December 2023.

**Supplementary Methods**

The used model for estimating infections and immunity was extended from a previously published model, *covidestim* [28], and later developments of that model [14, 15, 42]. Here, we describe the updated model and highlight the changes relative to the previously published model.

**Modeled disease states**

Figure 1C shows the modeled disease states and progression between states. A SARS-CoV-2 infection progresses, with assumed delays, from asymptomatic infection to symptomatic infection, to severe disease and eventually to death. At each disease state (except for death), individuals can recover and not progress to the next disease state. In each disease state, except for death, infected individuals can be diagnosed, which was modeled as a latent state. That is, an undiagnosed and diagnosed infection recover and progress with the same probability. All diagnosed cases (asymptomatic, symptomatic, severe) were assumed to be reported after a delay. We assumed a SARS-CoV-2 death is only reported if a diagnosis was made in a disease state before the death occurred. The modeled reported cases (sum of asymptomatic, symptomatic, and severe), modeled reported severe infections, and modeled reported deaths, were fit to the observed case, hospitalization, and death reports. Table S1 presents the original priors and delay distributions, and Table S2 presents the additional or altered assumptions.

**Progression probabilities and adjustment**

The progression probabilities each have a time-invariant prior distribution and have time-varying adjustments, that are further detailed in the remainder of this paragraph.

First, the probability to die if severely diseased is assumed to be increased in the beginning of the pandemic, when hospitals were overflowing, and health care was not readily accessible. This was modeled using a geographic region-specific adjustment, (*ifrAdjustment* Eq. S1).

Second, when vaccinations were developed, not all age groups were equally likely to be vaccinated. Specifically, older people received vaccinations before younger people did. Older people also have an assumed relative higher mortality rate from a SARS-CoV-2 infection. Together, this results in a reduced total mortality rate after infections when vaccinations are unequally taken up over age groups in the population. Because age is not included in the model, we accounted for this by calculating the relative risk reduction due to age specific vaccination imbalances outside of the model and dividing this adjustment over the three progression probabilities using a Dirichlet(3) prior (*D1, RRvaxAdjustment*, Eq. S1-S3).

Thirdly, prior infections and vaccinations give protection to progressing to severe disease for a subsequent infection. We calculate the fraction of the currently infected population with either no prior exposure or a prior exposure (vaccination, booster, infection) that is susceptible for infection *and* subsequent severe disease, and adjust the total probability to progress from infection to severe disease using a Dirichlet (2) prior. (*D2, RRsevProtAdjustment,* Eq. S2-S3).

Finally, the probability to develop symptomatic disease if infected is built up using a prior for the pre-Omicron time, a linear transition, and a prior for the since-Omicron time (*pSymIfInfAdj,* Eq. S3).

$pDieIfSev_{i} = pDieIfSev \times RRvaxAdjustment_{die,i}^D1(1) \times ifrAdjustment_{i}$	(Eq.S1)

$pSe{vIfSym}_{i}= pSevIfSym \times RRvaxAdjustment_{sev,i}^D1(2) \times RRsevProtAdjustment_{sev,i} ^D2(1)$	(Eq. S2)

$pSymIfInf_{i} = pSymIfInfAdj_{i} \times RRvaxAdjustment_{sym,i}^D1(3) \times RRsevProtAdjustment_{sym,i}^D2(2)$ (Eq. S3)

**Calculation of exposure state transitions and protection**

The model is constrained in the number of new infections at each time point by the susceptible population, which in turn is a function of the prevalence of exposure states and corresponding protection against infection in the population. Specifically, we consider four *exposure states*: naïve (never infected or vaccinated), infected (never vaccinated), vaccinated (never infected), and hybrid (at least one infection and at least one vaccination record) (Figure 1A). Each exposure state has an assumed protection against (re)infection and severe disease and subsequent waning of that protection. We use exponential waning of protection to make the algorithm memoryless, which is necessary for model fitting.

New infections can be *first infections* or *reinfections*, and the vaccination data separates out *first vaccination series* and *received booster doses.* These are assigned to the four exposure states in the following way. First, we assume that the new infections occur proportionally over the population susceptible to infection, accounting for prior acquired protection.

$newInf_{i,state} = infections_{i} \times(susInf_{i,state}/ susInf_{i})$	(Eq. S4)

Using Equation S4, new infections are assigned as *reinfections* in the *infected* or *hybrid* states, and *first infections* in the *naïve* and *vaccinated* states. Second, we assume that booster doses occur proportionally over *vaccinated* and *hybrid* states. Finally, we use the odds ratio of being vaccinated given prior infection, combined with the cumulative *first infections* and *vaccination series* to calculate the fraction of the total population *immunological exposed*, and subsequently determine the fraction of the population in the *hybrid* exposure state. The vaccinations are then assigned over the *naïve* and *infected* populations to match the required new *hybrid exposures*. If more hybrid exposures are required than vaccinations, the *first infection series* get redistributed over the *naïve* and *vaccinated* populations to correct this. Only in this situation, the infections are not distributed equally over the susceptible populations.

The prevalence in each exposure state is calculated by subtracting the exits and adding the entries into that state (Eq. S5). Reinfections and boosters are considered both an entry and an exit in the same state. This is necessary to calculate the protection appropriately.

The protection against infection and severe disease is calculated by multiplying the protection of the previous time with the fraction not exiting the exposure state, applying the exponential waning, and then adding the new entries (Eq. S6-7).

$Prvl_{<state>,i} = Prvl_{<state>,i-1}- exits_{<state>,i} + entries_{<state>,i}$	(Eq. S5)

$ProtInf_{<state>,i} =(ProtInf_{<state>,i-1} * (1-exi{ts}_{<state>,i}/Prvl_{<state>,i-1})) \times exp(waningInf_{<state>}) + entries_{<state>,i}$	(Eq. S6)

$ProtSev_{<state>,i} =(ProtSev_{<state>,i-1} * (1-exi{ts}_{<state>,i}/Prvl_{<state>,i-1})) \times exp(waningSev_{<state>})+ entries_{<state>,i}$	(Eq. S7)

**Vaccination data smoothing**

Our initial data source for COVID-19 vaccination data stopped reporting on May 13, 2023. We extended the vaccination data with monthly reported coverage of the updated 2023-2024 COVID-19 vaccination from the Immunization Information Systems, which was available from July 30, 2023 onward. We linearly disaggregated the monthly coverage to a weekly level, and we assumed no vaccinations were administered between May 13, 2023 and July 1, 2023. For seven states, no reported vaccination data were available from 2023 onwards from the Immunization Information Systems, and instead, weekly estimates from the National Immunization Survey were used.

| Model Parameter | Mean, std. Deviation | Distribution | Type | Source |
| --- | --- | --- | --- | --- |
| Log of New Infections At T=0 ($\boldsymbol{A}_{\boldsymbol{0}}$) | 0,10 | Normal(0,10) | prior | Assumed |
| $\boldsymbol{X}_{\boldsymbol{R,t}}$ Spline Parameters | 0,3 | Normal(0,3) | prior | Assumed |
| First Derivative of $\boldsymbol{X}_{\boldsymbol{R,t}}$Spline Parameters | 0,0.5 | Normal(0,0.5) | prior | Assumed |
| Second Derivative of $\boldsymbol{X}_{\boldsymbol{R,t}}$Spline Parameters | 0,0.1 | Normal(0,0.1) | prior | Assumed |
| Serial Interval | 5.8, 0.5 | Gamma(129.1, 22.25) | prior | [32] |
| Probability of Developing Symptoms If Infected | 0.59, 0.16 | Beta(5.14, 3.53) | prior | [33-35] |
| Probability of Becoming Severely Ill If Symptomatic | 0.09, 0.06 | Beta(1.89, 20.00) | prior | [37, 45] |
| Probability of Death for All Infections (national average) | 0.005, 0.001 | Beta(15.9, 3167) | prior | [30, 31] |
| Probability of Death for Severe Infections | 0.15, 0.03 | Beta(28.2, 162.3) | prior | [45] |
| Additional Risk of Mortality Prior to May 1 2020 ($\boldsymbol{a}_{\boldsymbol{p}_{\boldsymbol{D}}}$) | 1.34, 0.39 | Gamma(12.03, 8.99) | prior | Assumed |
| Rate Ratio, Diagnosis at Asymptomatic Vs. Symptomatic | 0.1, 0.07 | Beta(2,18) | prior | Assumed |
| Rate Ratio, Diagnosis at Symptomatic Vs. Severe | 0.5, 0.22 | Beta(2,2) | prior | Assumed |
| Probability of Diagnosis at Severe | 0.72, 0.16 | Beta(20,5) | prior | Assumed |
| Dispersion Parameter for Reported COVID-19 Cases (1/σ)^2^ | 0.8, 0.6 | Half-Normal(0,1) | prior | [29] |
| Dispersion Parameter for Reported COVID-19 Deaths (1/σ)^2^ | 0.8, 0.6 | Half-Normal(0,1) | prior | [29] |
| Scaling Factor: Time to Diagnosis Relative to Time in Symptomatic State | 0.5, 0.22 | Beta(2,2) | prior | Assumed |
| Scaling Factor: Time to Diagnosis Relative to Time in Severe State | 0.5, 0.22 | Beta(2,2) | prior | Assumed |

**Table S1: covidestim model parameters. Reproduced with permission from Chitwood et al., 2022.**

| **Parameter** | | **Description** | **Prior** |
| --- | --- | --- | --- |
| serial_i_pre | Length of the serial interval between infections prior to December 1, 2021 | | Gamma(129.1, 22.25)  ~ 5.5 days |
| serial_i_since | Length of the serial interval between infections since January 1, 2022 | | Gamma(34.615, 11.538*7)  ~ 0.4 weeks = 3 days |
| IFR_decl_OR | The increased infection fatality ratio in the early pandemic | | Gamma(12.031, 8.999) |
| p_sym_if_inf_pre | The probability of progressing to symptomatic disease if infected prior to December 1, 2021 | | Beta(5.1430, 3.5360) ~0.6 |
| p_sym_if_inf_since | The probability of progressing to symptomatic disease if infected after January 1, 2022 | | Beta(2.5,10) ~ 0.2 |
| p_sev_if_sym | The probability of progressing to severe disease if with symptomatic disease | | Beta(1.8854, 20.002)  ~0.08 |
| p_die_if_sev | The probability of progressing to death if with severe disease | | Beta(28.239, 162.30) |
| p_die_if_inf_pre | The probability of dying if infected, prior to December 1, 2022 | | Beta(15.915,3167.1)  ~.005 (national avg) |
| p_die_if_inf_since | The probability of dying if infected, after January 1, 2022 | | Beta(2.55,1594)  ~.001 |
| ifr_vac_adj | Dividing the total infection fatality ratio adjustment afforded by age differences in vaccination uptake over the transition probabilities p_sym_if_inf_(pre/since), p_sev_if_sym, and p_die_if_sev | | Dirichlet(3) |
| isr_prot_adj | Dividing the total infection severe ratio adjustment afforded by protection against severe disease from prior immune-exposure (infection, vaccination or both) over the transition probabilities p_sym_if_inf and p_sev_if_sym | | Dirichlet(2) |

**Table S2: Additional covidestim model parameters and prior distributions.**

| Location | Date of maximum protection against SARS-CoV-2 infection | Maximum protection against SARS-CoV-2 infection  [95%CrI] | December 2023 protection against SARS-CoV-2 infection [95%CrI] |
| --- | --- | --- | --- |
| United States | 2023-02-25 | 53.6 [38.7–71.5] | 55.3 [43.1–68.7] |
| U.S. Census regions | | | |
| Northeast  (excluding Vermont) | 2022-02-25 | 57.6 [40.8–72.5] | 60.7 [45.6–73.7] |
| West | 2023-12-30 | 56.3 [41.8–74.2] | 56.3 [41.8–74.2] |
| South | 2023-12-30 | 52.1 [37.2–70.2] | 55.8 [43.6–69.8] |
| Midwest | 2023-02-19 | 50.1 [35.9–71.5] | 51.3 [46.1–56.0] |
| us states | | | |
| Alabama | 2023-12-30 | 56.2 [33.6–72.9] | 56.2 [33.6–72.9] |
| Alaska | 2022-09-10 | 47.1 [31.4–67.4] | 55.9 [43.6–65.0] |
| Arizona | 2023-01-21 | 61.3 [43.7–74.1] | 65.4 [51.4–74.6] |
| Arkansas | 2023-01-21 | 57.7 [37.7–72.7] | 59.2 [44.5–73.3] |
| California | 2023-12-30 | 54.1 [41.5–74.2] | 54.1 [41.5–74.2] |
| Colorado | 2023-12-30 | 56.3 [40.7–76.4] | 56.3 [40.7–76.4] |
| Connecticut | 2023-03-04 | 59.8 [40.2–73.0] | 62.4 [44.5–74.5] |
| Delaware | 2023-02-25 | 60.2 [42.4–71.7] | 67.6 [49.4–76.0] |
| District Of Columbia | 2022-02-19 | 38.4 [29.4–50.4] | 47.9 [43.3–53.3] |
| Florida | 2023-09-30 | 55.8 [40.0–70.8] | 57.3 [41.1–72.2] |
| Georgia | 2022-02-19 | 41.6 [29.3–66.3] | 52.2 [46.0–58.6] |
| Hawaii | 2023-12-02 | 61.1 [46.2–71.9] | 61.4 [46.6–72.5] |
| Idaho | 2023-12-30 | 56.4 [33.1–72.7] | 56.4 [33.1–72.7] |
| Illinois | 2022-02-19 | 47.6 [36.4–66.2] | 51.5 [46.7–57.0] |
| Indiana | 2022-02-19 | 43.1 [29.9–66.7] | 51.3 [44.6–56.4] |
| Iowa | 2023-12-30 | 55.3 [35.0–73.7] | 55.3 [35.0–73.7] |
| Kansas | 2023-12-30 | 59.0 [42.2–74.4] | 59.0 [42.2–74.4] |
| Kentucky | 2023-02-25 | 53.0 [36.1–70.3] | 58.3 [43.0–73.5] |
| Louisiana | 2022-03-19 | 47.7 [32.2–70.0] | 47.9 [43.8–52.4] |
| Maine | 2023-03-25 | 67.7 [49.5–75.0] | 72.0 [53.1–76.0] |
| Maryland | 2023-02-18 | 51.6 [39.1–73.1] | 59.3 [48.5–76.0] |
| Massachusetts | 2023-12-30 | 67.1 [41.6–76.1] | 67.1 [41.6–76.1] |
| Michigan | 2023-12-30 | 53.5 [35.9–72.4] | 53.5 [35.9–72.4] |
| Minnesota | 2023-12-30 | 52.7 [40.4–74.3] | 52.7 [40.4–74.3] |
| Mississippi | 2022-02-26 | 36.1 [24.8–60.7] | 53.8 [45.3–63.3] |
| Missouri | 2022-02-19 | 47.7 [33.5–71.1] | 49.0 [43.2–54.4] |
| Montana | 2022-03-05 | 47.0 [29.6–70.5] | 48.4 [40.7–53.9] |
| Nebraska | 2023-12-30 | 54.9 [38.4–71.2] | 54.9 [38.4–71.2] |
| Nevada | 2022-08-13 | 55.4 [36.3–73.7] | 61.4 [49.4–73.4] |
| New Hampshire | 2023-12-30 | 53.7 [43.2–70.3] | 53.7 [43.2–70.3] |
| New Jersey | 2023-02-18 | 60.3 [43.5–73.3] | 68.2 [51.4–76.2] |
| New Mexico | 2022-12-17 | 66.7 [60.5–72.5] | 75.7 [71.6–76.4] |
| New York | 2023-02-04 | 52.9 [37.6–71.0] | 58.4 [45.2–74.5] |
| North Carolina | 2023-02-18 | 54.6 [39.2–76.1] | 59.6 [45.6–75.0] |
| North Dakota | 2022-02-19 | 49.9 [36.1–71.4] | 50.6 [45.5–55.1] |
| Ohio | 2022-02-12 | 48.2 [35.8–69.3] | 51.8 [46.4–57.0] |
| Oklahoma | 2023-02-18 | 59.1 [41.7–71.8] | 65.1 [49.3–74.7] |
| Oregon | 2023-12-30 | 65.8 [46.4–76.1] | 65.8 [46.4–76.1] |
| Pennsylvania | 2023-03-11 | 56.8 [42.3–72.3] | 58.8 [46.5–73.1] |
| Rhode Island | 2023-12-30 | 52.2 [39.4–68.2] | 52.2 [39.4–68.2] |
| South Carolina | 2023-02-25 | 58.4 [40.7–72.7] | 59.8 [45.0–72.4] |
| South Dakota | 2023-12-30 | 57.8 [41.6–71.1] | 57.8 [41.6–71.1] |
| Tennessee | 2023-03-04 | 59.0 [38.7–71.8] | 60.6 [45.2–72.4] |
| Texas | 2022-09-17 | 47.9 [36.2–66.8] | 54.2 [46.4–65.4] |
| Utah | 2023-12-30 | 58.3 [42.1–75.5] | 58.3 [42.1–75.5] |
| Virginia | 2023-02-25 | 57.2 [44.3–74.3] | 57.5 [45.2–71.5] |
| Washington | 2023-12-30 | 56.9 [41.9–74.3] | 56.9 [41.9–74.3] |
| West Virginia | 2023-12-30 | 54.4 [37.2–74.6] | 54.4 [37.2–74.6] |
| Wisconsin | 2022-02-19 | 51.0 [36.3–70.7] | 53.2 [47.9–57.4] |
| Wyoming | 2022-02-26 | 46.4 [28.6–68.4] | 51.6 [43.4–57.6] |

**Table S3: Summary of immunological protection for the United States, U.S. Census Regions, U.S. States and the District of Columbia, 2020-2023.**

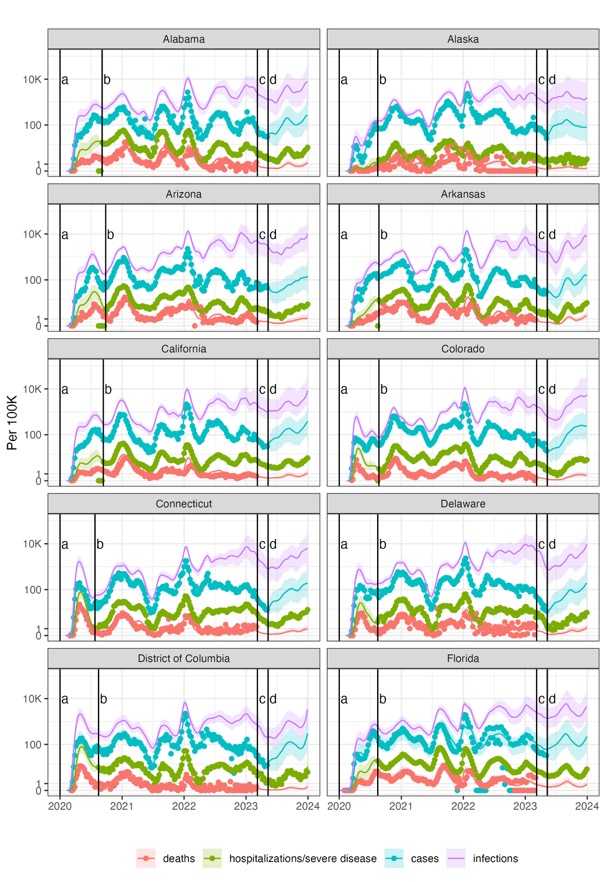

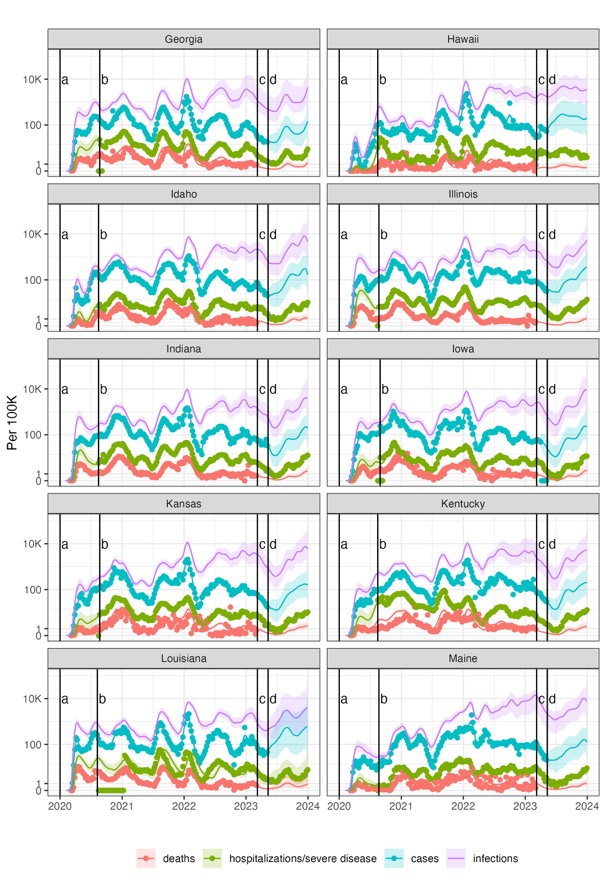

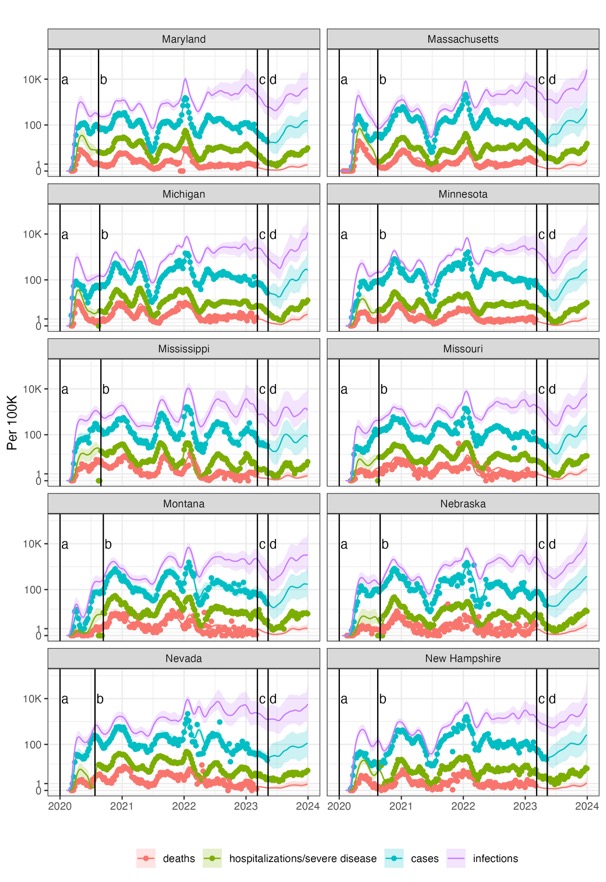

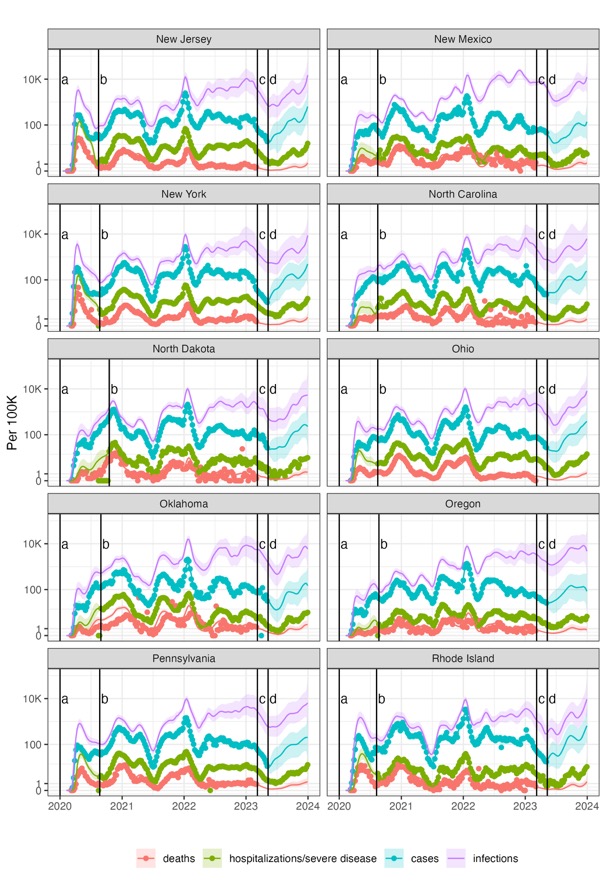

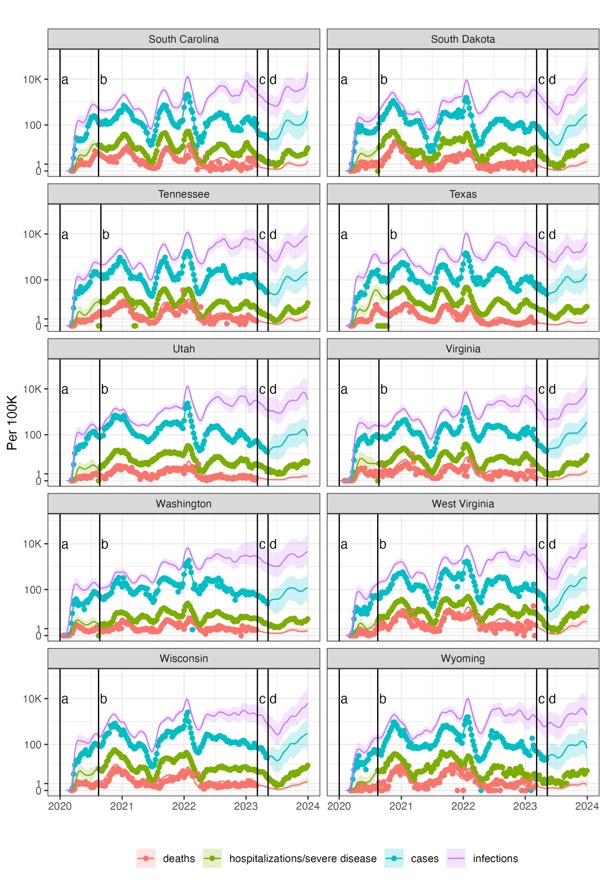

**Figure S1: Model estimated SARS-CoV-2 infections and COVID-19 disease outcomes compared with reported data, for each U.S. state and District of Columbia, January 2020–December 2023.**

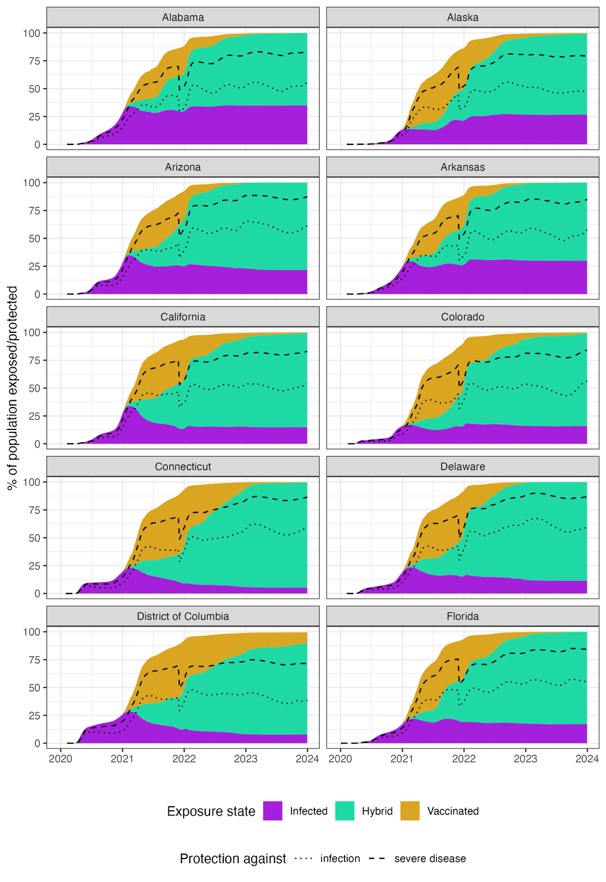

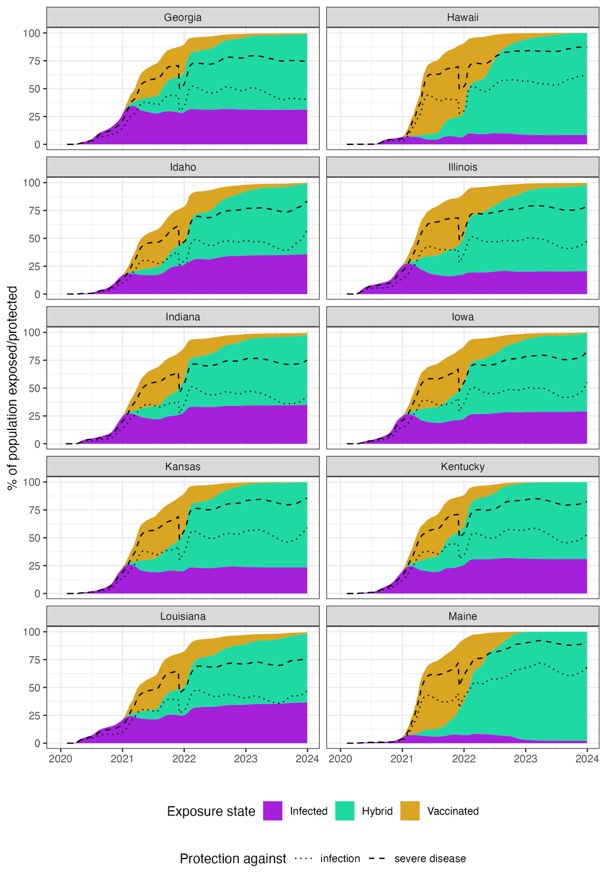

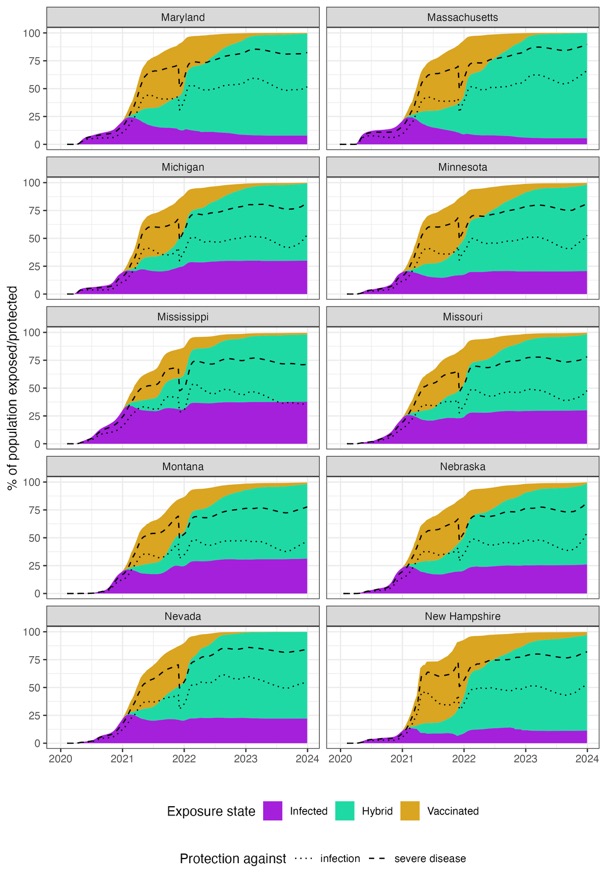

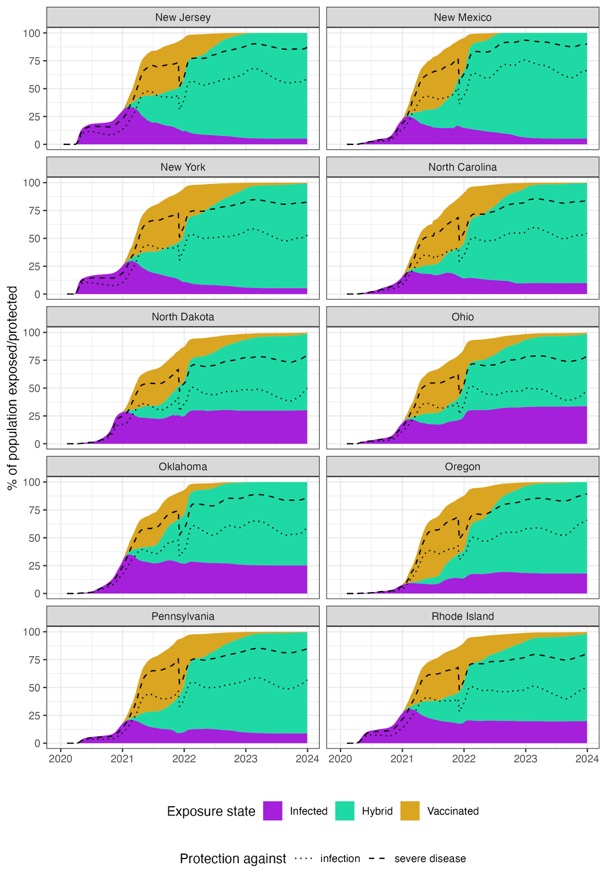

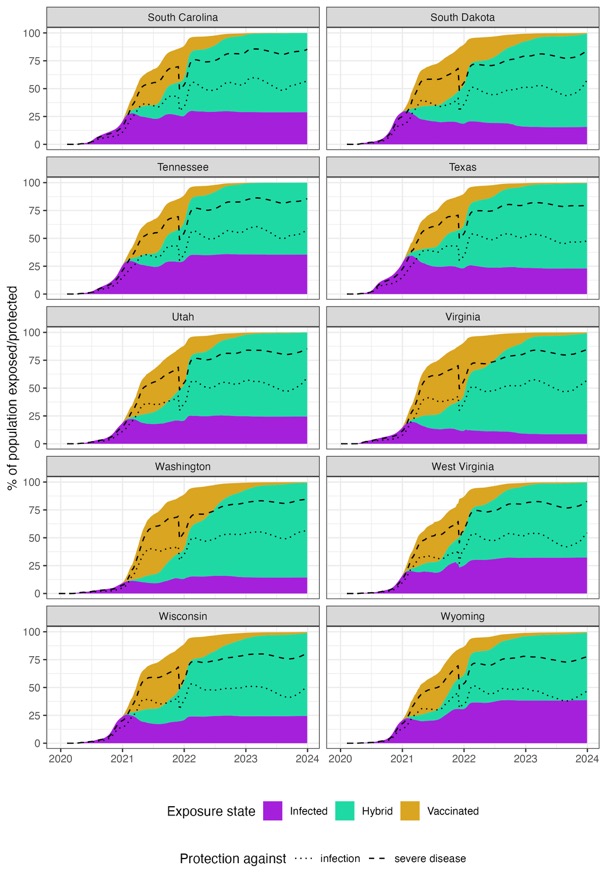

**Figure S2: Percentage of population immunological exposed and protected against SARS-CoV-2 infection and severe COVID-19 disease stratified exposure route and U.S. state, January 2020 – December 2023.**

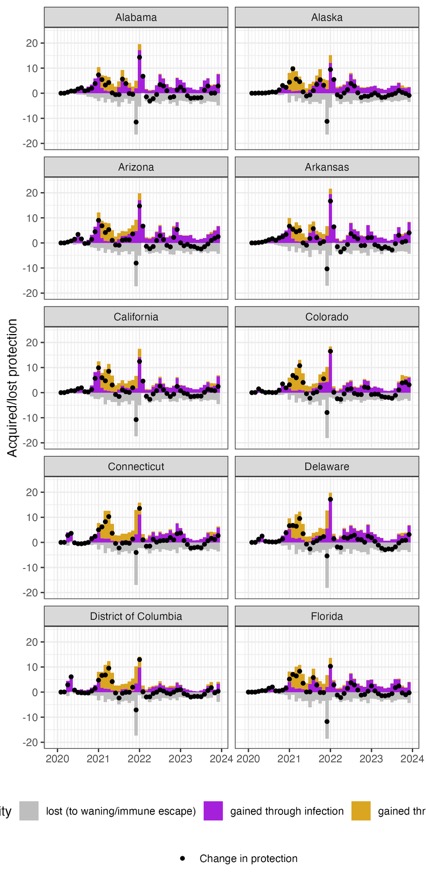

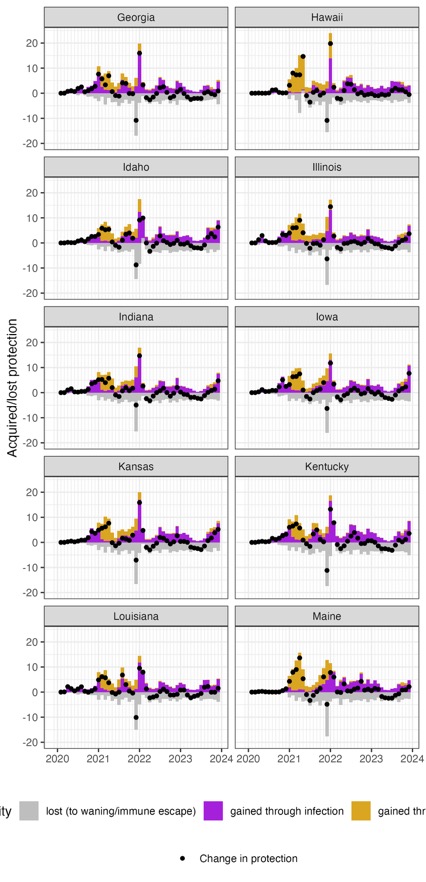

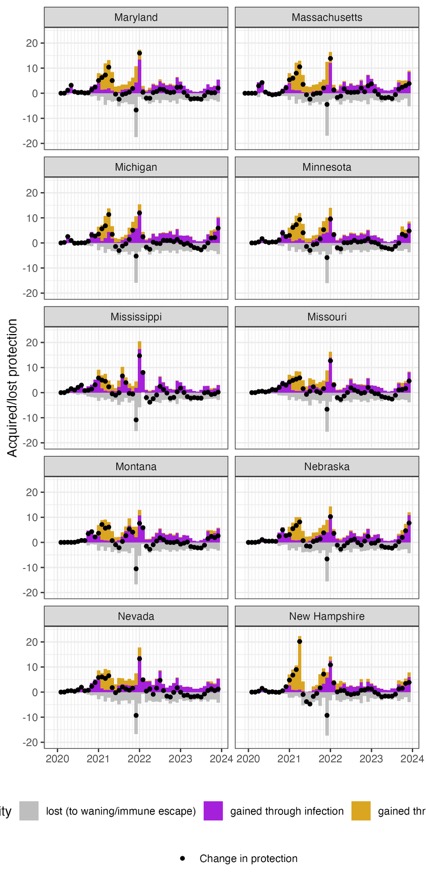

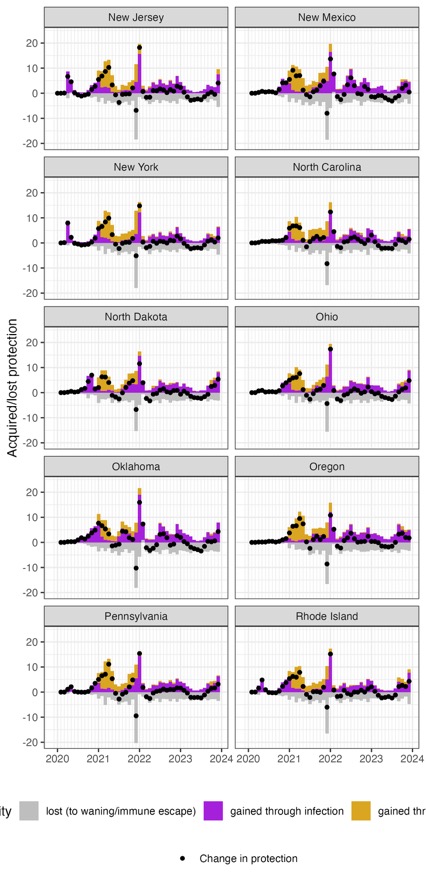

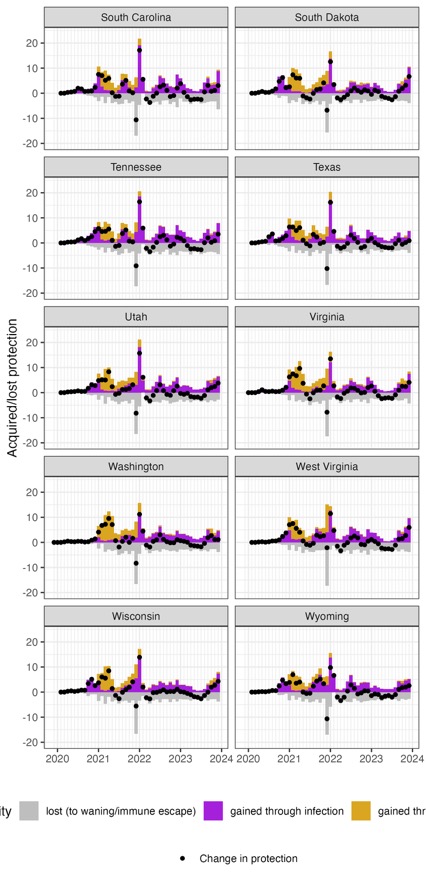

**Figure S3: Monthly change in acquired immunity stratified by exposure route and U.S. state, 2020–2023.**

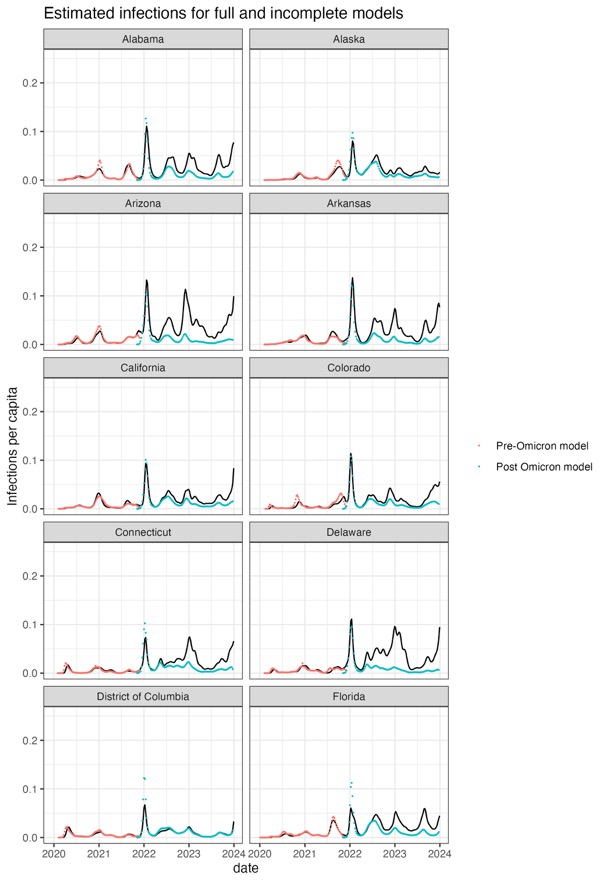

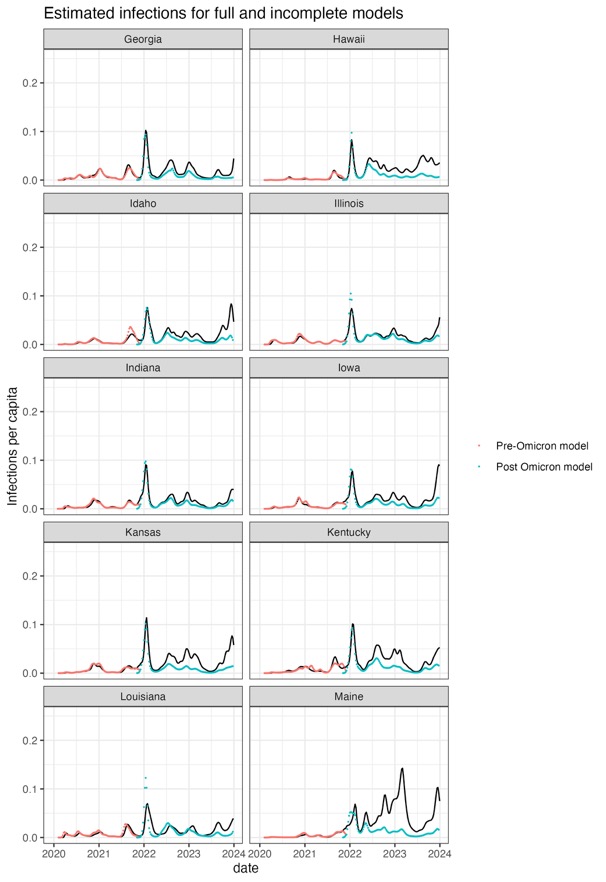

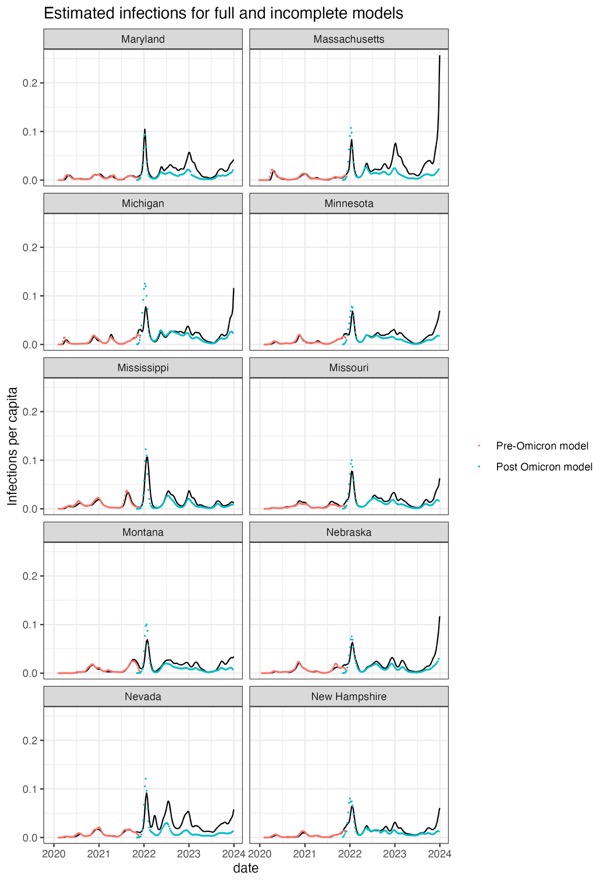

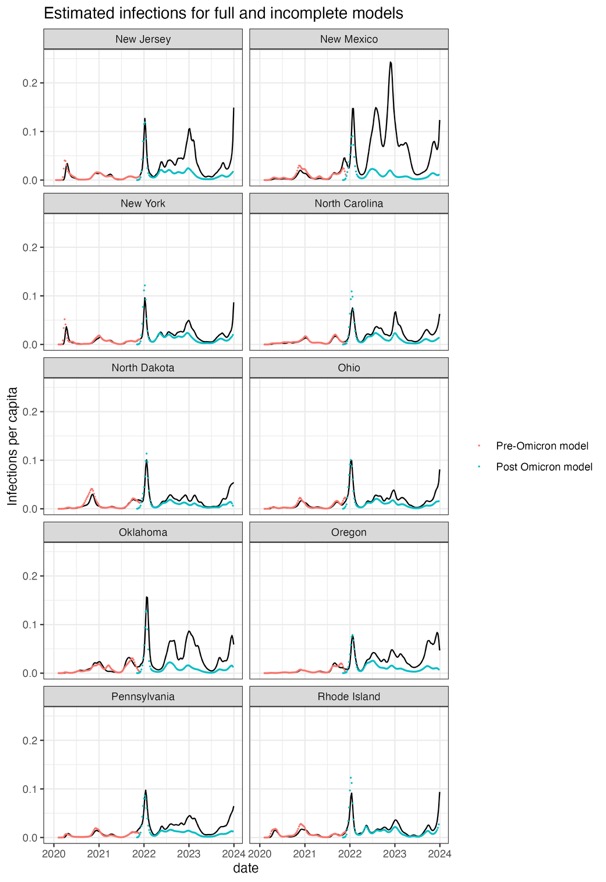

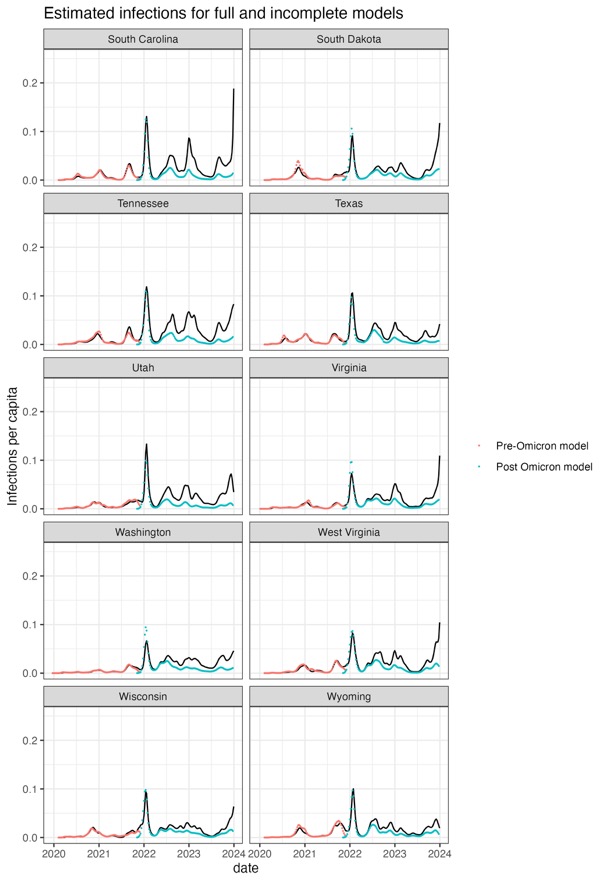

**Figure S4: Comparison of SARS-CoV-2 infection estimates updated *covidestim* model versus earlier divided pre- and post-Omicron model estimates.**

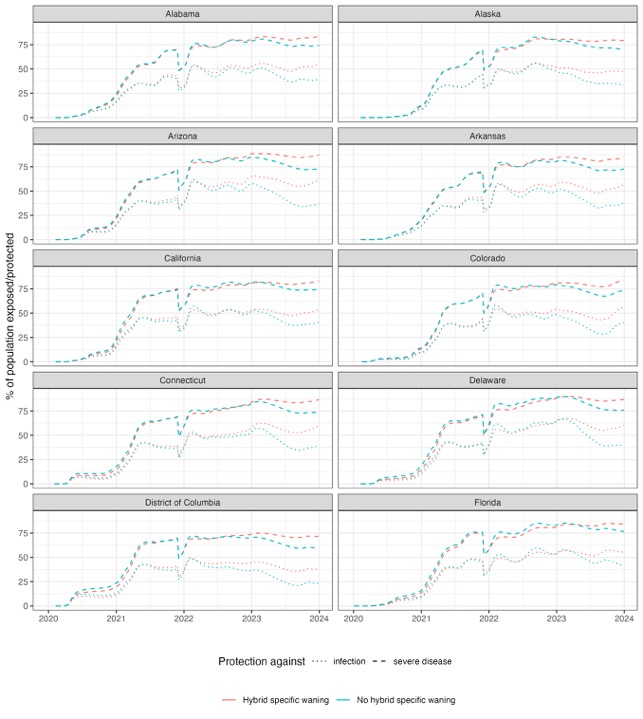

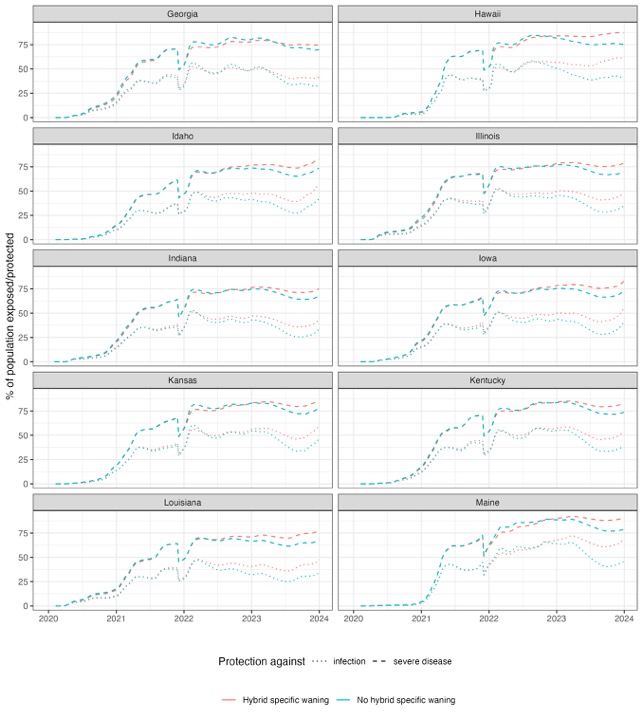

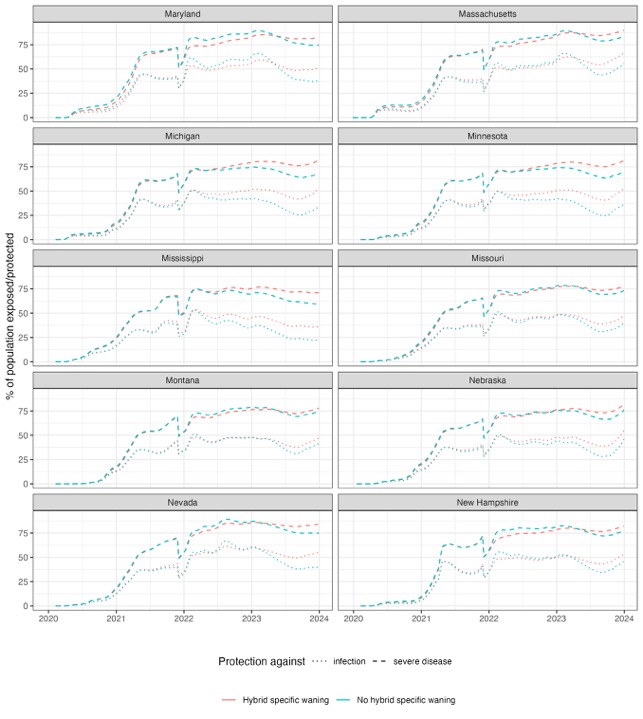

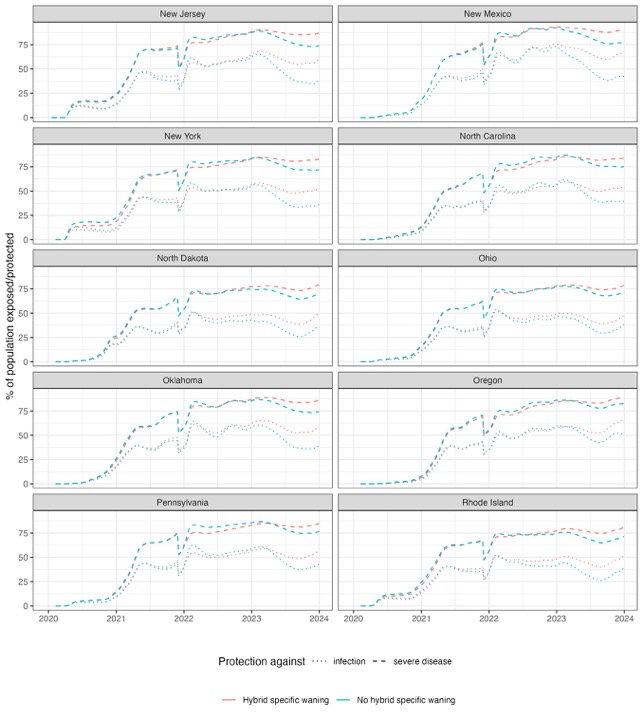

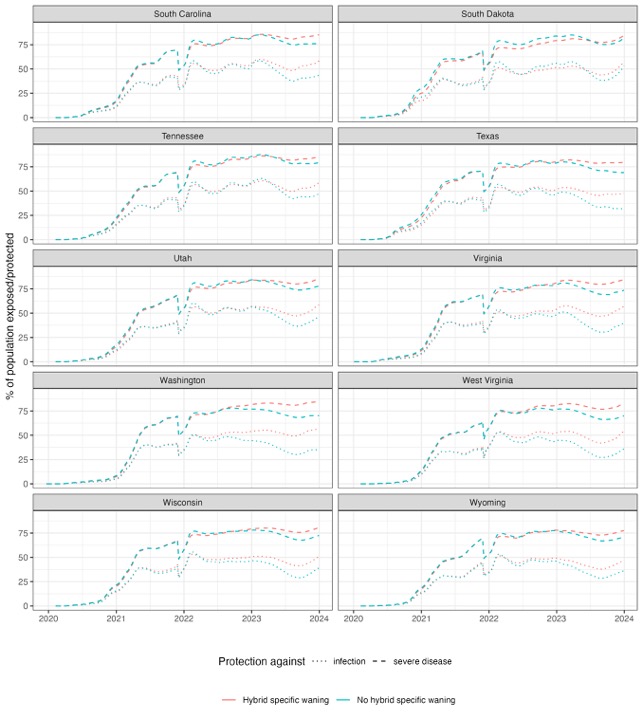

**Figure S5: Comparison of effective protection against SARS-CoV-2 infection and severe COVID-19 disease with and without hybrid immunity specific waning assumptions, for each U.S. state and the District of Columbia, January 2020–December 2023.**
